# Physician Composition of Hospitals’ Workforce And Mortality Across U.S. Hospitals

**DOI:** 10.1101/2025.10.31.25339262

**Authors:** Cameron J. Gettel, Zhuohui Lin, Craig Rothenberg, Zhenqiu Lin, Tara Lagu, Kate Goodrich, Joseph S. Ross, Arjun K. Venkatesh

## Abstract

**Importance:** The clinical workforce composition in U.S. hospitals is shifting, with advanced practice practitioners (APPs) – nurse practitioners and physician assistants –assuming larger roles in inpatient care. How this mix relates to patient mortality is unclear.

**Objective:** To investigate the association between hospital-level physician proportion and 30-day risk standardized mortality rate (RSMRs) for six common inpatient conditions.

**Design:** Cross-sectional national study linking 2022 CMS Physician & Other Practitioners, Facility Affiliation, Hospital Compare Complications and Deaths, and American Hospital Association Annual Survey data. Analyses were completed October 17, 2025.

**Setting:** Nation-wide U.S. hospitals treating traditional Medicare beneficiaries in 2022.

**Participants:** Medicare beneficiaries hospitalized for acute myocardial infarction, Chronic Obstructive Pulmonary Disease (COPD), Coronary Artery Bypass Graft (CABG) Surgery, heart failure, pneumonia, or stroke.

**Exposure(s) (for observational studies):** Hospital-level physician proportion, defined as the number of physicians divided by the sum of affiliated physicians and APPs.

**Main Outcome(s) and Measure(s):** Hospital-level condition-specific 30-day RSMRs – case-mix adjusted outcome measures presented as proportions.

**Results:** Among 3,487 hospitals (mean physician proportion 79.7% [SD, 9.4%]), mean physician proportions across quartiles ranged from 67.8% (Q1; n=872; range, 25.0–73.5%) to 91.6% (Q4; n=871; range, 86.7–99.6%). Mean hospital-level RSMRs (%, 95% CI) were 12.63 (12.57–12.69) for acute myocardial infarction, 2.93 (2.88–2.99) for CABG surgery, 9.50 (9.44–9.56) for COPD, 12.03 (11.96–12.11) for heart failure, 18.12 (18.02–18.21) for pneumonia, and 13.74 (13.66–13.82) for stroke. Hospitals in the highest physician proportion quartile (Q4) had lower RSMRs than hospitals [p value, 95% CI] in the lowest quartile (Q1) for all conditions – acute myocardial infarction (12.54 vs. 12.86 [p<0.001, 0.16-0.48]), CABG (2.92 vs. 3.05 [p=0.106, -0.29-0.30]), COPD (9.11 vs. 9.83 [p<0.001, 0.56-0.88]), heart failure (11.24 vs. 12.73 [p<0.001, 1.29-1.70]), pneumonia (17.51 vs. 18.64 [p<0.001, 0.87-1.39]), and stroke (13.41 vs. 14.31 [p<0.001, 0.67-1.12]). Generalized additive models identified significant non-linear associations between the physician proportion and the RSMRs for COPD, acute myocardial infarction, heart failure, pneumonia, and stroke, respectively explaining 4.77-11.82% of the deviance.

**Conclusions and Relevance:** Higher relative physician staffing was modestly but consistently associated with lower hospital-level mortality across common and high-burden medical conditions. Workforce composition may be a key structural determinant of hospital quality and warrants consideration in workforce and quality improvement strategies.

## Introduction

The composition of the healthcare workforce in U.S. hospitals is undergoing a significant transformation. Over the past two decades, the number of advanced practice practitioners (APPs) – including nurse practitioners (NPs) and physician assistants (PAs) – has risen dramatically to address physician shortages and expand care access.^1–3^ APPs frequently serve as physician extenders in hospital medicine, critical care, and surgical services – managing routine floor care, coordinating discharges, and conducting peri-operative assessments. This shift is particularly salient in hospitals that face persistent challenges in recruiting or retaining physicians, where APPs may be deployed more broadly and often practice in settings with limited access to specialty consultation or support.^4^

There is long-standing evidence linking hospital structure and process characteristics to patient outcomes, with the clinical workforce representing a particularly influential component. A substantial body of research has demonstrated associations between nurse staffing ratios, nurse education and experience, and key outcomes such as in-hospital mortality.^5,6^ While prior literature has additionally explored the influence of hospital characteristics on patient mortality,^7–9^ relatively little is known about how the clinician workforce composition may contribute to variation in these outcomes. Much of the existing empirical work related to APPs has focused on ambulatory or primary care settings, or on process-oriented outcomes such as imaging utilization or healthcare spending.^10–13^ Fewer studies have examined how the relative mix of physicians and APPs within hospitals may influence clinical outcomes, particularly for acutely ill or complex patients.

Understanding how the composition of the clinical workforce – physicians and APPs – is associated with hospital performance has important implications. As financial pressures in healthcare delivery and value-based care models are increasingly promulgated, APPs are increasingly being integrated into accountable care organizations and other value-based payment models. Yet there is limited evidence on how these expanded roles affect clinical outcomes – particularly mortality – within hospital settings. If greater reliance on APPs is associated with different patient outcomes, this has critical implications not only for workforce planning and scope-of-practice regulations, but also for health system accountability under quality-based reimbursement. In this context, we aimed to evaluate whether the proportion of physicians within a hospital’s clinician workforce is associated with risk standardized mortality rates for six common and high-burden medical conditions. We hypothesized that hospitals with a higher proportion of physicians would demonstrate lower mortality rates, potentially reflecting differences in training, decision-making capacity, or hospital-wide team structure.

## Methods

### Data Sources and Study Population

This cross-sectional study included hospitalizations of traditional Medicare, aged 65 and older, treated by physicians and APPs. The study was deemed exempt by the Human Investigation Committee at the Yale University School of Medicine, New Haven, Connecticut. The Strengthening the Reporting of Observational Studies in Epidemiology reporting guideline for cross-sectional studies was followed.

To identify eligible hospitals and their associated clinical workforce, we began by linking National Provider Identifier (NPI)-level data of practicing clinicians from the 2022 Centers for Medicare & Medicaid Services (CMS) Physician & Other Practitioners dataset (n=1,230,293 unique NPIs)^14^ with the CMS Doctors and Clinicians Facility Affiliation dataset (n=775,288 unique NPIs at hospitals).^15^ The Facility Affiliation dataset, which provides links between individual clinicians and the hospitals where they provided Medicare services, was used to identify hospitals via CMS Certification Numbers (CCNs).

The identified NPIs were then grouped into corresponding hospitals (n=4,381 unique CCNs) they were affiliated with (Figure 1). These hospitals were then linked to the CMS Hospital Compare Complications and Deaths – Hospital dataset (n=4,779 unique CCNs)^16^, which provided hospital-level quality performance via risk-standardized mortality rates (RSMRs) of six conditions/procedures of interest: coronary artery bypass graft (CABG) surgery, chronic obstructive pulmonary disease (COPD), acute myocardial infarction, heart failure, pneumonia, and stroke. Hospital quality has commonly been evaluated using RSMRs,^17–23^ which account for differences in patient demographics and comorbidities between hospitals to allow fair comparisons across institutions. Hospitals without RSMR data or located outside the 50 states and DC were excluded (Figure 1). Finally, linkable by CCN, we obtained characteristics of included hospitals (Table 1) using the 2022 American Hospital Association (AHA) Annual Survey (n=5,921 unique CCNs), resulting in 3,487 hospitals eligible for the analysis. These datasets have been previously used in workforce analyses.^24–26^

**Figure 1.**
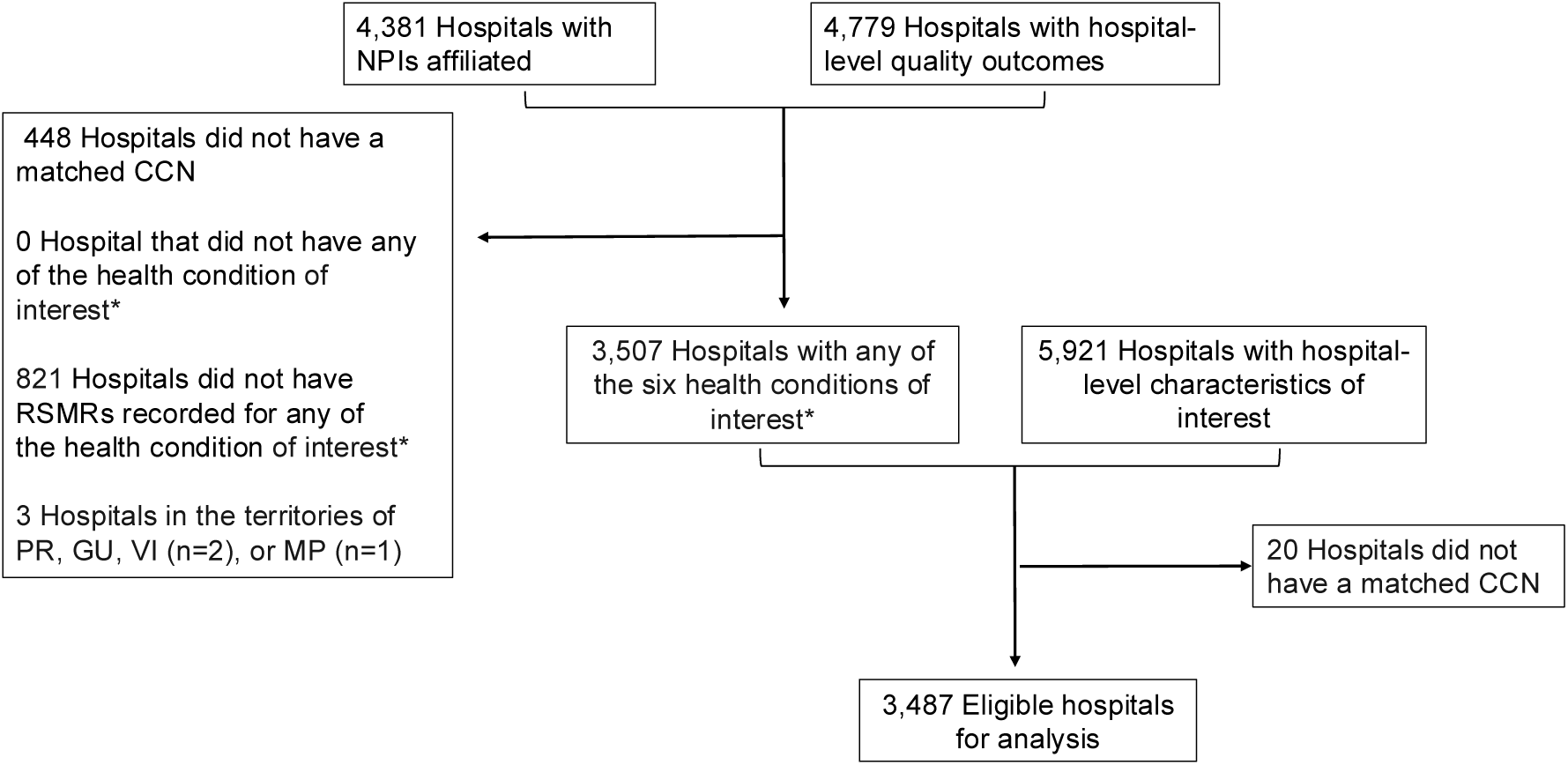
Derivation of the analytic cohort. Abbreviation: NPI, National Provider Identifier; CCN, CMS Certification Numbers; RSMR: 30-day risk-standardized mortality rate; PR: Puerto Rico; VI: U.S. Virgin Islands; GU: Guam; MP: Commonwealth of the Northern Mariana Islands. * Indicates COPD, CABG surgery, pneumonia, stroke, acute myocardial infarction, heart failure.

**Table 1.**
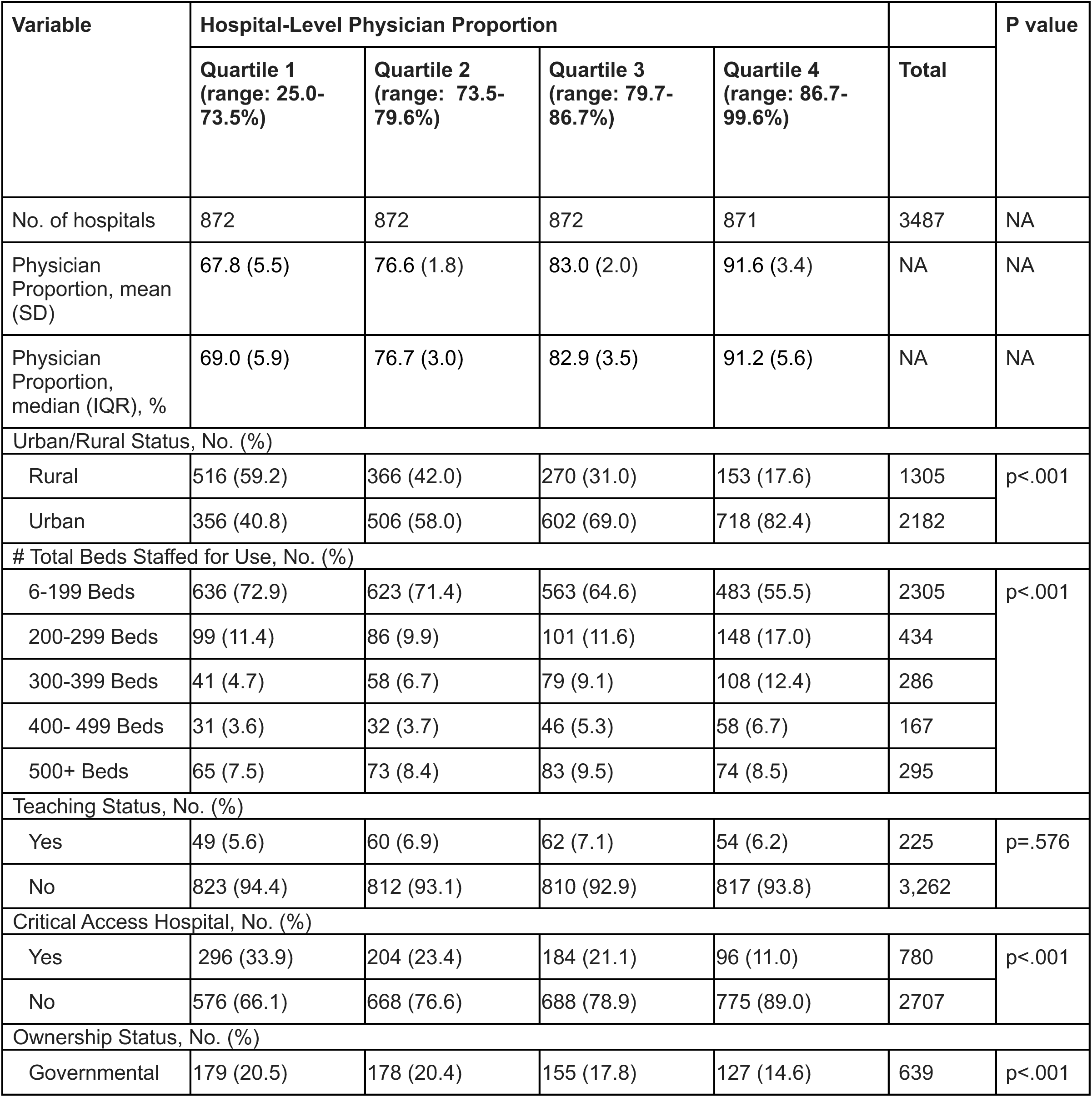
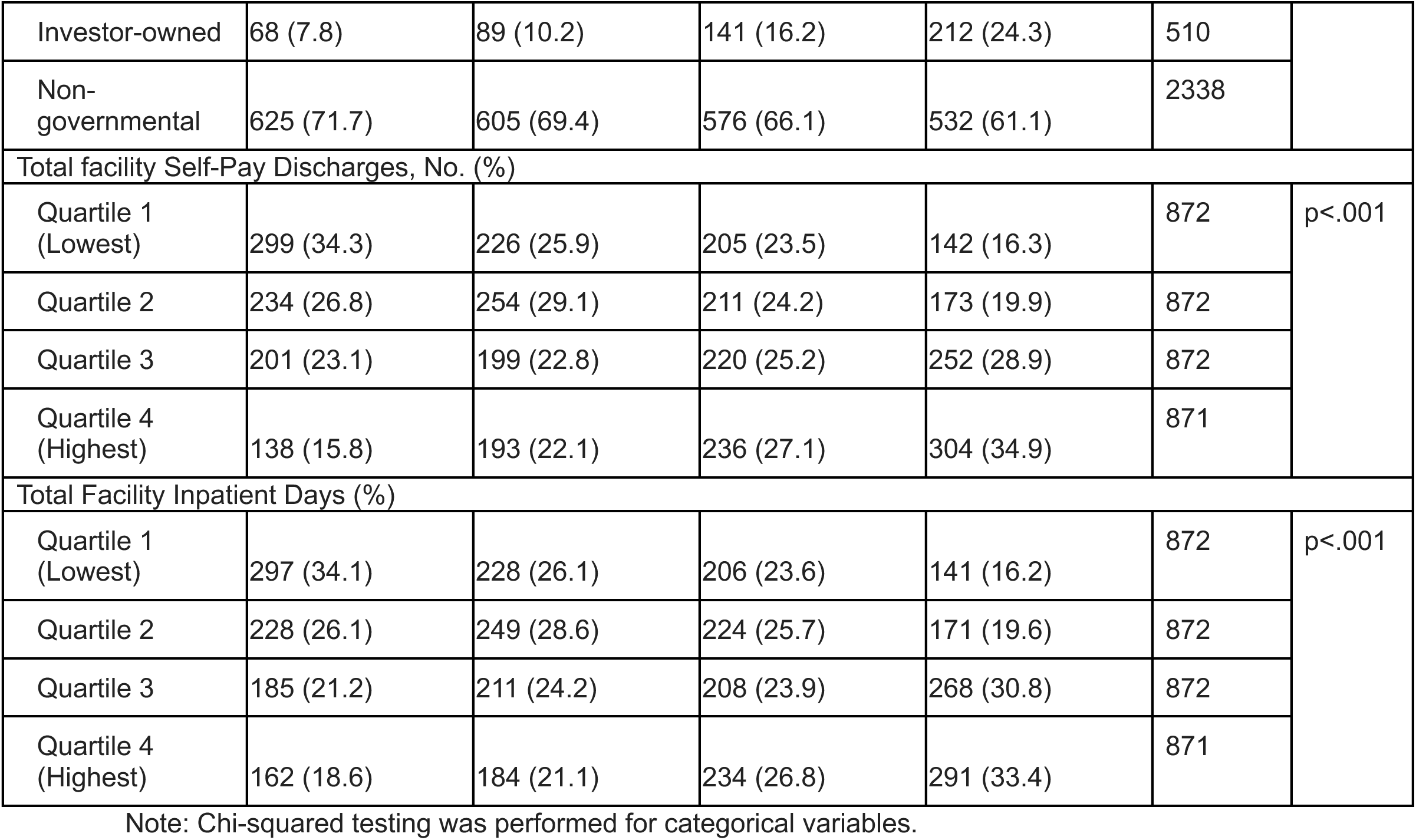
Characteristics of hospitals, stratified by physician proportion.

Within the analytic sample, physicians were defined using specialty keywords of “MD” (Doctor of Medicine) and “DO” (Doctor of Osteopathic Medicine), while advanced practice practitioners (APPs) were defined based on specialty keywords “Nurse Practitioner” and “Physician Assistant”.

### Hospital Characteristics

Available hospital characteristics included urban/rural status, the number of beds regularly staffed for use (6-199, 200-299, 300-399, 400-499, and 500+), teaching status (Yes/No), critical access hospital status (Yes/No), ownership status (government [including non-federal and federal], for-profit [e.g. investor-owned], and non-profit). In addition, several continuous hospital characteristic variables – total facility self-pay discharges and total facility inpatient days – were categorized into quartiles with approximately equal numbers of hospitals (Q1: lowest, Q4: highest) to summarize variation across hospitals and facilitate descriptive comparisons in baseline characteristics. Hospitals were divided into quartiles based on their physician proportion using a rank-based method, which assigns approximately equal numbers of hospitals to each quartile.

### Statistical Analysis

The primary exposure was the proportion of a hospital’s workforce comprised of physicians, calculated by dividing the number of physicians affiliated with the hospital by the combined number of physicians and APPs affiliated with the hospital (eFigure 1). We initially grouped hospitals by the physician proportion into quartiles to characterize and compare hospital characteristics using analysis of variance for continuous variables and χ² tests for categorical variables. We further compared the mean RSMR difference between Quartile 1 and Quartile 4 of physician proportion for each condition using Welch’s two-sided t test.

At the level of the hospital, we then examined associations between the physician proportion and the primary outcome – RSMRs of the six conditions of interest. RSMRs – presented as proportion – are case-mix adjusted outcome measures that reflect 30-day mortality for specific conditions, allowing for standardized comparisons of hospital performance across institutions regardless of differences in patient populations. Calculated Pearson correlation coefficients were weighted by the number of inpatient beds, serving as a proxy for hospital volume. To estimate 95% confidence intervals for the weighted correlations, we used a nonparametric bootstrap method with 1,000 iterations. The Pearson coefficient ranges from –1 to +1, with negative values indicating an inverse association (i.e., higher physician proportion associated with lower RSMRs), positive values indicating a direct association, and values near zero indicating little to no linear relationship.

To assess potential nonlinear relationships, we then fitted generalized additive (GAM) models with cubic regression splines to model the association between physician proportion and RSMR for each diagnosis. To account for both patient- and hospital-level factors, the models were adjusted for staffing ratio (total inpatient hospital beds divided by hospital physician counts; see eFigure 2) and urban/rural status and were weighted by hospital volume. Predictions were generated across the observed range of physician proportion for each diagnosis, holding the staffing ratio constant at its median value and including both rural and urban hospitals. This aimed to isolate the marginal effect of physician proportion on RSMR while preserving the inherent complexity of the hospital system. The 95% confidence intervals were computed based on the standard errors of the fitted values. The proportion of variance in RSMR explained by physician proportion (i.e., the shared variance) was quantified using the deviance explained from each GAM. This approach was aligned with Desai et al.’s assessment of the variation in the value of care for acute myocardial infarction, heart failure, and pneumonia.^27^ In this analysis, the RSMR was treated as the dependent variable and the hospital’s physician proportion as the continuous independent variable. Analyses were conducted using R software (version 2024.04.2). Weighted correlations were computed using the wCorr package, models were fitted using the mgcv package, and data visualizations were generated using ggplot2.

## Results

### Sample Characteristics

Among 4,381 hospitals with NPIs affiliated and 4,779 hospitals with hospital-level quality outcomes, 3,487 hospitals met eligibility criteria were included in the analysis (Figure 1). Among 3,487 hospitals with publicly reported hospital mortality rates in 2022, the average physician proportion was 79.7% (SD: 9.4) (eFigure 1). Of these, 3,449 hospitals had available data for pneumonia RSMRs, 2,984 for heart failure, 2,506 for COPD, 2,193 for stroke, 1,898 for acute myocardial infarction, and 887 for CABG surgery (Table 1). Hospitals varied substantially in structural characteristics. Overall, 62.5% of hospitals were located in urban areas, 66.1% had fewer than 200 staffed beds, and 22% were designated critical access hospitals. Most were non-governmental (67%), and 6.4% were teaching hospitals.

### Clinician Workforce Composition and Hospital Characteristics

Hospitals in Quartile 1, representing the lowest physician proportion (25.0–73.5%), had a median (IQR) and mean (SD) physician proportion of 69.0% (5.9) and 67.8% (5.5), respectively. In contrast, hospitals in Quartile 4, which had the highest physician proportion (86.7–99.6%), had a median and mean of 91.2% (5.6) and 91.6% (3.4), respectively. Hospitals with higher physician proportions differed from those with lower proportions. Compared to Quartile 1 hospitals, those in Quartile 4 were significantly more likely to be located in urban areas (82.4% vs. 40.8%), to have larger bed counts (≥300 beds in 27.6% of hospitals vs. 15.8%), and to report greater total inpatient days and higher proportions of self-pay discharges. Quartile 4 hospitals were also less likely to be critical access hospitals (11.0% vs. 33.9%) (Table 1).

### Mortality Rates by Physician Proportion Quartile

Hospitals with higher physician proportions exhibited lower risk-standardized mortality rates across all six conditions studied (Table 2). Hospitals in the highest physician proportion quartile (Q4) had lower RSMRs than hospitals [p value, 95% CI] in the lowest quartile (Q1) for all conditions – acute myocardial infarction (12.54 vs. 12.86 [p<0.001, 0.16-0.48]), CABG (2.92 vs. 3.05 [p=0.106, -0.29-0.30]), COPD (9.11 vs. 9.83 [p<0.001, 0.56-0.88]), heart failure (11.24 vs. 12.73 [p<0.001, 1.29-1.70]), pneumonia (17.51 vs. 18.64 [p<0.001, 0.87-1.39]), and stroke (13.41 vs. 14.31 [p<0.001, 0.67-1.12]).

**Table 2.** Hospital-level 30-day risk-standardized mortality rates for six conditions, stratified by physician proportion.

| Diagnostic condition | No. of hospitals | Risk-standardized mortality rates, mean (SD, 95% CI), % |  |  |  |  | P value |
| --- | --- | --- | --- | --- | --- | --- | --- |
|  |  | Overall | Physician Proportion Q1 | Physician Proportion Q2 | Physician Proportion Q3 | Physician Proportion Q4 |  |
| Acute Myocardial Infarction | 1898 | 12.63 (1.28, 12.57-12.69) | 12.86 (1.35, 12.73-12.99) | 12.65 (1.37, 12.52-12.77) | 12.54 (1.25, 12.43-12.65) | 12.54 (1.15, 12.44-12.64) | p<.001 |
| CABG Surgery | 887 | 2.93 (0.83, 2.88-2.99) | 3.05 (0.93, 2.93-3.18) | 2.96 (0.81, 2.86-3.07) | 2.81 (0.81, 2.71-2.91) | 2.92 (0.75, 2.82-3.02) | p=.017 |
| COPD | 2506 | 9.50 (1.49, 9.44-9.56) | 9.83 (1.54, 9.70-9.96) | 9.66 (1.49, 9.55-9.78) | 9.43 (1.55, 9.31-9.55) | 9.11 (1.27, 9.01-9.21) | p<.001 |
| Heart failure | 2984 | 12.03 (2.08, 11.96-12.11) | 12.73 (2.00, 12.58-12.88) | 12.45 (2.10, 12.30-12.60) | 11.81 (2.02, 11.67-11.96) | 11.24 (1.87, 11.10-11.37) | p<.001 |
| Pneumonia | 3449 | 18.12 (2.80, 18.02-18.21) | 18.64 (2.84, 18.45-18.82) | 18.35 (2.86, 18.16-18.55) | 17.97 (2.73, 17.78-18.15) | 17.51 (2.66, 17.33-17.69) | p<.001 |
| Stroke | 2193 | 13.74 (1.87, 13.66-13.82) | 14.31 (1.88, 14.14-14.48) | 13.85 (1.92, 13.69-14.01) | 13.53 (1.77, 13.38-13.68) | 13.41 (1.82, 13.27-13.55) | p<.001 |

Summary statistics (mean, SD, 95%CI) and sample size were provided. ANOVA was performed for continuous variables. Hospital sample sizes for the RSMRs of the six conditions of interest reflects the number of hospitals with publicly available mortality data for that condition. Note: As an example interpretation, for heart failure, the mean RSMRs differed significantly across quartiles of physician proportion (ANOVA, $p < 0.001$ ). Hospitals in the lowest quartile (25.0– 73.5%) had the highest mean RSMR (12.73%, 95% CI: 12.58% -12.88%), whereas hospitals in highest quartiles (86.7-99.6%) had the lowest RSMRs (11.24%, 95% CI: 11.10%-11.37%).

### Correlation and Model-Based Associations

In correlation analyses, we identified inverse associations between the physician proportion and RSMRs for 5 of the 6 conditions of interest weighting by inpatient beds – – acute myocardial infarction, COPD, heart failure, pneumonia, and stroke – and a non-significant positive association for CABG surgery (-0.059, 95% CI: -0.134 to 0.014). The strength of the association was modest, and varied by condition, with respective Pearson correlation coefficients (r) of -0.080 (95%CI, -0.139 to -0.017) for acute myocardial infarction, -0.172 (95%CI, -0.219 to -0.120) for COPD, -0.275 (95%CI, -0.327 to -0.220) for heart failure, -0.105 (95%CI, -0.156 to -0.055) for pneumonia, and -0.149 (95%CI, -0.202 to -0.093) for stroke. To complement the overall correlation analyses, we also examined whether the association between physician proportion and RSMRs varied across quartiles of physician proportion weighted by hospital volume. For each quartile, we calculated disease-specific weighted Pearson correlation, and the findings indicated that the correlations did not display a consistent pattern of linearly increasing or decreasing strength across quartiles (Table 3).

**Table 3.** Pearson correlation of hospital physician proportion and risk-standardized mortality rates weighted by hospital volume.

| Physician Proportion Quartile | Acute Myocardial Infarction |  | CABG Surgery |  | COPD |  | Heart Failure |  | Pneumonia |  | Stroke |  |
| --- | --- | --- | --- | --- | --- | --- | --- | --- | --- | --- | --- | --- |
|  | No. of hospital | Correlation (95% CI) | No. of hospital | Correlation (95% CI) | No. of hospital | Correlation (95% CI) | No. of hospital | Correlation (95% CI) | No. of hospital | Correlation (95% CI) | No. of hospital | Correlation (95% CI) |
| Quartile 1 (Lowest) | 398 | -0.111 (-0.236 to 0.023) | 209 | 0.048 (-0.070 to 0.153) | 569 | -0.119 (-0.230 to -0.013) | 675 | -0.081 (-0.237 to 0.056) | 862 | -0.058 (-0.161 to 0.041) | 469 | -0.006 (-0.116 to 0.117) |
| Quartile 2 | 450 | 0.026 (-0.108 to 0.159) | 221 | -0.071 (-0.219 to 0.092) | 650 | -0.033 (-0.153 to 0.090) | 762 | -0.108 (-0.216 to -0.005) | 865 | -0.026 (-0.133 to 0.089) | 549 | -0.043 (-0.156 to 0.073) |
| Quartile 3 | 505 | -0.084 (-0.218 to 0.051) | 246 | -0.169 (-0.307 to -0.028) | 643 | -0.080 (-0.199 to 0.038) | 774 | -0.109 (-0.207 to -0.007) | 863 | -0.076 (-0.182 to 0.037) | 561 | -0.087 (-0.211 to 0.039) |
| Quartile 4 (Highest) | 545 | -0.072 (-0.166 to 0.028) | 211 | 0.036 (-0.097 to 0.176) | 644 | 0.062 (-0.037 to 0.157) | 773 | -0.049 (-0.141 to 0.040) | 859 | 0.023 (-0.071 to 0.119) | 614 | -0.024 (-0.114 to 0.065) |

To reduce confounding by key structural factors while preserving the complexity inherent to hospital systems, particularly the uneven distribution of urban and rural hospitals across quartiles (Table 1) and differences in hospital volume per clinician, we employed generalized additive models, adjusting for rural versus urban status and staffing ratio, and weighting by overall hospital volume. The findings showed consistent trend with quartile-based, disease-specific correlation, where there were significant non-linear associations between the physician proportion and the RSMRs for COPD, acute myocardial infarction, heart failure, pneumonia, and stroke (Figure 2). The deviance explained by the models was 4.77%, 6.5%, 5.41%, 11.82%, 8.22%, and 6.06% for acute myocardial infarction, CABG surgery, COPD, heart failure, pneumonia, and stroke, respectively.

**Figure 2.**
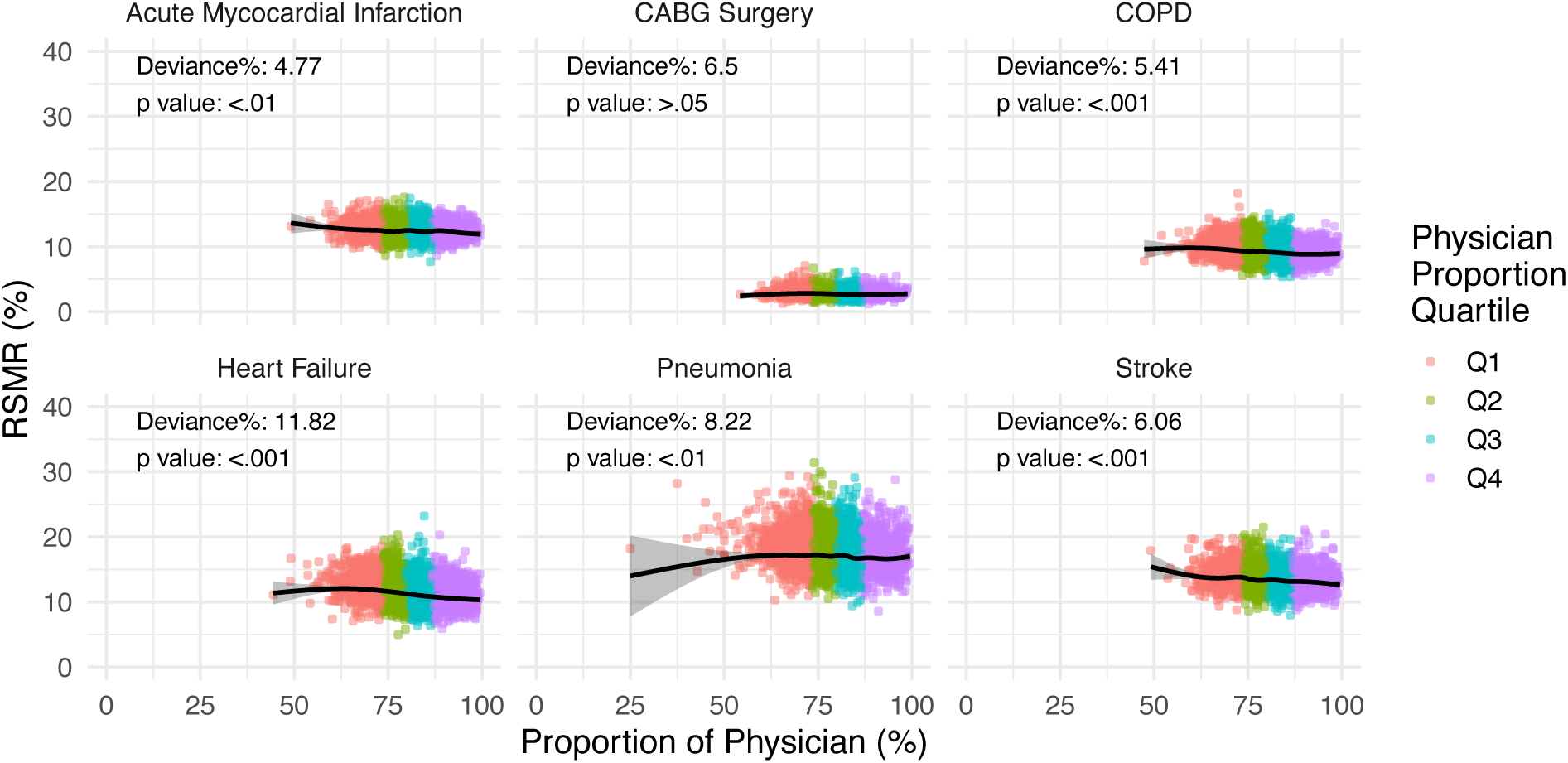
Scatterplot of hospital-level physician proportion and risk-standardized mortality rates for six conditions. Black lines show the cubic spline smooth regression lines with risk-standardized mortality rates (RSMRs) as the dependent variable and proportion of physician as the independent variable. The shadowed areas around the cubic spline regression lines show the 95% CIs. Colored dots indicate individual hospitals by quartile of physician proportion. Percent deviance explained and p-values are shown for each condition. Note – As an example interpretation, 11.82% of the variation in hospital-level heart failure risk-standardized mortality rate is explained by physician proportion. The p value of .001 indicates the physician proportion is significantly non-linearly associated with RSMR.

## Discussion

In this large, cross-sectional analysis of 3,487 U.S. hospitals, we found that a higher proportion of physicians relative to advanced practice practitioners providing hospital care was modestly but consistently associated with lower hospital-level risk-standardized mortality rates for several common conditions: CABG, COPD, acute myocardial infarction, heart failure, pneumonia, and stroke. These associations were statistically significant and held after weighting for staffing ratio and adjusting for urban/rural status, suggesting that workforce composition may constitute an important quality structure that contributes to variation in hospital mortality across these common and high-impact conditions. The mechanisms underlying this association are likely complex and multifactorial.

Prior research has demonstrated that APPs can provide care comparable to physicians in a range of ambulatory settings, including chronic disease management, preventive care, and routine primary care.^30,32,33^ Similarly, in hospital-based studies, APPs have been shown to perform well within integrated care teams – particularly in institutions with strong physician oversight, well-defined protocols, and stable patient populations.^34,35^ However, most of these studies have focused on individual-level encounters, process-based outcomes or low-acuity patient populations. Our study adds to this discourse by examining hospital-level outcomes rather than individual-level encounters, highlighting a modest but consistent association between greater relative physician representation and improved mortality performance for conditions that frequently require intensive, multidisciplinary management.

The strength and pattern of associations across conditions suggest that physician-led or physician-directed care may be particularly impactful in managing complex, high-acuity diagnoses – and may also serve as a proxy for broader institutional capacity, such as access to specialty consultation and advanced diagnostic capabilities that support high-quality care delivery. The strongest inverse association was observed for heart failure (r = -0.275), a condition that often requires complex clinical decision-making, nuanced medication management, and close coordination between hospital and outpatient providers. Similar associations were identified for COPD and stroke – conditions that also demand timely diagnosis and intervention and evidence-based management. Although the absolute effect sizes were modest, these findings support the hypothesis that hospitals with a greater proportion of physicians as part of their workforce may have structural or clinical advantages that translate into improved patient outcomes. Interestingly, we found no statistically significant association between physician proportion and RSMRs for CABG surgery during correlation analyses. This may reflect the fact that surgical outcomes are heavily influenced by the experience of individual surgeons, surgical team dynamics, and perioperative infrastructure – factors that may not correlate closely with general hospital workforce composition.

Rather than interpreting these results as evidence of superior individual performance by physicians, our findings may reflect broader institutional patterns. That is, hospitals with a higher proportion of physicians may differ systematically from those with greater APP representation in ways that are not fully captured by observed characteristics. These differences could include variations in case complexity, staffing models, care coordination infrastructure, or access to specialty services. Therefore, we caution against overinterpreting these differences. The cross-sectional nature of our analysis limits causal inference, and the ecologic fallacy is a relevant concern; we do not know whether patients with worse outcomes were cared for by APPs, nor do we know the precise roles, responsibilities, or supervision structures of APPs across hospitals. The relationship we observed is at the hospital level and does not reflect individual clinician performance or patient-level assignment of care. However, the global hospital-level findings do imply that policies and staffing decisions should focus less on simplistic clinician substitution and more on ensuring appropriate deployment, integration, and support of diverse clinical roles within hospital care teams.

Our study provides valuable insights into the relationship between workforce composition and hospital quality performance. As health systems continue to evolve and adapt to workforce shortages, value-based payment reform, and growing patient complexity, our findings underscore the importance of evaluating how care team structures affect clinical outcomes. Clinician mix decisions should balance cost, staffing, and quality. Payers increasingly emphasize performance on outcome and utilization metrics in contracting and network design decisions; clinical workforce composition as a structural indicator of quality could prove useful to payers and providers alike. Furthermore, newer alternative payment models should be designed to support optimal team structures without creating economic incentives or cost metrics that carry the unintended consequence of favoring fewer physician complements and in turn lower quality. Health system leaders, policymakers, and accrediting bodies may even consider incorporating workforce composition metrics into quality improvement frameworks and staffing guidelines. Future research should explore the mechanisms underlying these associations, including how team-based models of care can be optimized to leverage the strengths of both physicians and APPs. Additionally, longitudinal studies and natural experiments, such as those induced by staffing changes or policy shifts, may offer deeper insight into the causal impact of workforce configuration on patient outcomes.

## Limitations

There are limitations to this study. First, the cross-sectional design precludes causal inference; we cannot determine whether increasing physician proportion would directly reduce mortality rates. Second, our data were limited traditional Medicare aged 65 and older, which may not generalize to younger populations or those enrolled in Medicare Advantage or other insurance plans. Third, clinician counts reflect hospital-wide affiliations rather than condition-specific clinical teams; while this is the best available national data linking clinicians to hospitals, it may not capture the precise mix of providers regularly managing patients with conditions such as COPD or stroke on a day-to-day basis. Additionally, a small number of hospitals had low total counts of affiliated clinicians (e.g., fewer than five), meaning that their physician proportion could be influenced by small denominators; however, such hospitals represented a minority of the analytic sample and are unlikely to have driven the overall associations observed. Fourth, hospital participation in CMS-reported mortality measures is limited to facilities with at least 25 qualifying index hospitalizations per condition, which excludes some small or low-volume hospitals from the analysis; for example, fewer hospitals reported CABG mortality, limiting power. Finally, hospitals with a greater proportion of physicians were more likely to be larger, urban, and less likely to be designated as critical access hospitals. Although we adjusted for hospital volume and categorized several key institutional characteristics, unmeasured confounding (e.g. nurse-to-patient ratios, the presence of hospitalists or intensivists, or the availability of clinical decision support tools) remains possible.

## Conclusion

In conclusion, hospitals with a higher proportion of physicians were modestly associated with lower risk-standardized mortality rates for several major conditions. These findings suggest that clinician workforce composition may be an important and under-recognized structure of quality that is essential to advancing hospital quality improvement efforts and patient outcomes. Optimizing care delivery will likely require not only the right mix of clinicians, but also strong systems of integration, supervision, and support.

## Data Availability

All data produced in the present study are available upon reasonable request to the authors

https://data.cms.gov/provider-summary-by-type-of-service/medicare-physician-other-practitioners/medicare-physician-other-practitioners-by-provider-and-service

https://data.cms.gov/provider-data/dataset/27ea-46a8

https://data.cms.gov/provider-data/topics/hospitals

## Authors Contributions

Dr. Gettel and Zhuohui Lin designed the study. Zhuohui Lin managed the data. Dr. Gettel, Zhuohui Lin, Craig Rothenberg, Dr. Lin, Dr. Venkatesh provided statistical advice on study design and analyzed the data. Dr. Gettel and Zhuohui Lin drafted the manuscript, and all authors contributed substantially to its revision. Dr. Gettel and Zhuohui Lin takes responsibility for the paper as a whole.

## Source of Funding

Dr. Gettel is supported by the National Institute on Aging (NIA) of the National Institutes of Health (R03AG073988) and the National Academy of Medicine of the National Academy of Sciences under award number SCON-10000824. The funders had no role in the design and conduct of the study; collection, management, analysis, and interpretation of the data; and preparation or approval of the manuscript.

## Conflicts of Interest

Dr. Gettel, Dr. Lin, and Dr. Venkatesh receive support for contracted work from the Centers for Medicare and Medicaid Services to develop hospital and healthcare outcome and efficiency quality measures. Dr. Lagu is an employee of Alliant Insurance Services; Alliant had no role in any aspect of this work. Dr. Ross currently receives research support through Yale University from Johnson and Johnson to develop methods of clinical trial data sharing, from the Food and Drug Administration for the Yale-Mayo Clinic Center for Excellence in Regulatory Science and Innovation (CERSI) program (U01FD005938), from the NIH, from the Greenwall Foundation, and from Arnold Ventures.

## Notes

### Author Declarations

Secondary data analysis deemed exempt by institutional review board

