## Supplemental Figure 1 & 2 for "Physician Composition of Hospitals’ Workforce And Mortality Across U.S. Hospitals"

**Supplemental Files**

**eFigure 1. Hospital-level physician proportion across 3,487 hospitals**

Frequency distribution of hospitals by physician proportion (x-axis) and number of hospitals (y-axis). Note: physician proportion is defined as the number of affiliated physicians divided by the total number of affiliated physicians and APPs.

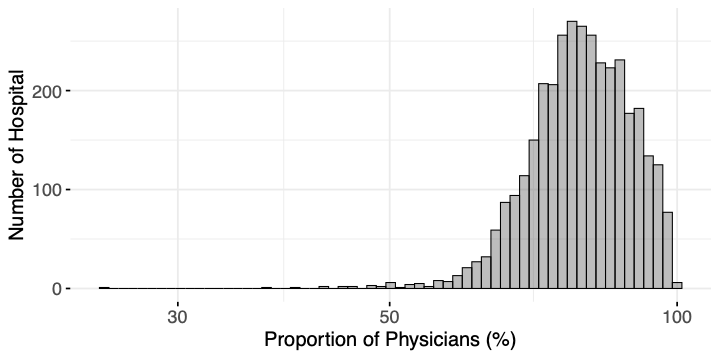

**eFigure 2. Histogram of staffing ratio across 3487 hospitals.**

Frequency distribution of hospitals by staffing ratio (x-axis) and number of hospitals (y-axis). Note: staffing ratio defined as the total inpatient hospital beds divided by hospital physician counts.

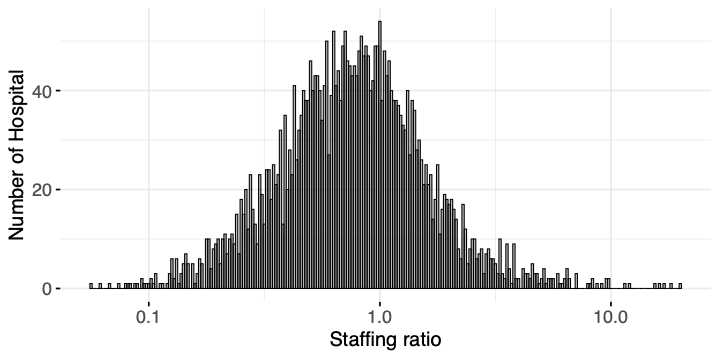
